# Executive function network’s white matter alterations relate to Parkinson’s disease motor phenotype

**DOI:** 10.1101/2020.08.15.20174284

**Authors:** Qinglu Yang, Shruti Nanivadekar, Paul A. Taylor, Zulin Dou, Codrin Lungu, Silvina G. Horovitz

## Abstract

Parkinson’s disease (PD) patients with postural instability and gait disorder phenotype (PIGD) are at high risk of cognitive deficits compared to those with tremor dominant phenotype (TD). Alterations of white matter (WM) integrity can occur in patients with normal cognitive functions (PD-N). However, the alterations of WM integrity related to cognitive functions in PD-N, especially in these two motor phenotypes, remain unclear. Diffusion tensor imaging (DTI) is a non-invasive neuroimaging method to evaluate WM properties and by applying DTI tractography, one can identify WM tracts connecting functional regions. Here, we 1) compared the executive function (EF) in PIGD phenotype with normal cognitive functions (PIGD-N) and TD phenotype with normal cognitive functions (TD-N) phenotypes; 2) used DTI tractography to evaluated differences in WM alterations between these two phenotypes within a task-based functional network; and 3) examined the WM integrity alterations related to EF in a whole brain network for PD-N patients regardless of phenotypes.

Thirty-four idiopathic PD-N patients were classified into two groups based on phenotypes: TD-N and PIGD-N, using an algorithm based on UPDRS part III. Neuropsychological tests were used to evaluate patients’ EF, including the Trail making test part A and B, the Stroop color naming, the Stroop word naming, the Stroop color-word interference task, as well as the FAS verbal fluency task and the animal category fluency tasks. DTI measures were calculated among WM regions associated with the verbal fluency network defined from previous task fMRI studies, and compared between PIGD-N and TD-N groups. In addition, the relationship of DTI measures and verbal fluency scores were evaluated for our full cohort of PD-N patients within the whole brain network. These values were also correlated with the scores of the FAS verbal fluency task.

Only the FAS verbal fluency test showed significant group differences, having lower scores in PIGD-N when compared to TD-N phenotype (*p* < 0.05). Compared to the TD-N, PIGD-N group exhibited significantly higher MD and RD in the tracts connecting the left superior temporal gyrus and left insula, and those connecting the right pars opercularis and right insula. Moreover, compared to TD-N, PIGD-N group had significantly higher RD in the tracts connecting right pars opercularis and right pars triangularis, and the tracts connecting right inferior temporal gyrus and right middle temporal gyrus. For the entire PD-N cohort, FAS verbal fluency scores positively correlated with MD in the superior longitudinal fasciculus (SLF).

This study confirmed that PIGD-N phenotype has more deficits in verbal fluency task than TD-N phenotype. Additionally, our findings suggest: (1) PIGD-N shows more microstructural changes related to FAS verbal fluency task when compared to TD-N phenotype; (2) SLF plays an important role in FAS verbal fluency task in PD-N patients regardless of motor phenotypes.

## 1. Introduction

Parkinson’s disease (PD) is a major neurodegenerative disorder, with a central feature of neurodegeneration of dopaminergic neurons in the substantia nigra pars compacta [1,2]. The most common manifestations include the classic triad of motor symptoms of rest tremor, rigidity, and bradykinesia. PD patients can be classified into two groups with distinct motor phenotypes: 1) tremor-dominant (TD), and 2) being postural instability and gait difficulty (PIGD) [3,4]. Non-motor symptoms, such as olfactory and cognitive decline related to PD, can occur 5 to 10 years before motor symptoms onset [5,6], suggesting that non-motor symptoms could be prodromal signs. Cognitive impairment, developing in some cases to dementia, is the most reported non-motor PD symptom affecting patients’ quality of life [7].

Recent studies investigating the relationship between motor and cognitive symptoms in PD suggest that motor phenotype is an important influential factor affecting cognitive function [8,9]. The PIGD phenotype has been correlated with increased risk for dementia [8–11]. In a longitudinal study [8], TD phenotype with normal cognitive function (TD-N) at the first time point (yrs. of disease duration: mean=9.5, SD=5.6) showed progression to dementia only after patients transitioned to PIGD. This transition was associated with accelerated cognitive decline and highly increased risk for subsequent dementia. PIGD phenotype, with normal cognitive functions (PIGD-N) at the first time point (yrs. of disease duration: mean=6.9, SD=4.7), remained within their phenotype and had a significant higher odds ratio for dementia compared to those in TD. Another study following nondemented PD patients for 2 years showed similar results [11]. These reports suggest cognitive impairment pathology may share neuro-anatomical or neuro-chemical substrates with a specific motor phenotype, and encourage us to further explore the underpinnings of the cognitive decline in the different PD motor phenotypes.

It has been proposed that alterations in white matter (WM) integrity might influence cognitive functions in PD [12,13]. Deterioration of WM microstructure correlates with cognitive impairment in PD across different cognitive stages, from mild impairment to dementia [12]. Neuroimaging studies showed FA reductions in the superior longitudinal fasciculus (SLF) in the PIGD-N phenotype and increased MD in the motor circuits in the TD-N phenotype [14]. Further exploring these differences might unveil additional information about the mechanisms of how motor phenotypes relate to cognitive function in PD.

Diffusion tensor imaging (DTI) is a non-invasive imaging method to study the microstructure of WM integrity *in vivo* [15]. Three commonly used measurements of DTI are fractional anisotropy (FA), mean diffusivity (MD), and radial diffusivity (RD) [16]. Using probabilistic tractography, it is possible to estimate locations of WM bundles, and then to examine changes in diffusion parameters within each tract [17]. Such changes may help us to understand the pathological processes underlying diseases [18]. In addition, WM alterations have been observed before gray matter (GM) atrophy in PD [19], and has been suggested they might help assessing cognitive impairment early in the process of cognitive decline in PD.

In this study, we used DTI probabilistic tractography, as well as cognitive and clinical data from a cohort of PD-N to better understand: 1) the effects of motor phenotypes in executive function; 2) the effects of motor phenotypes in WM properties within a task-based executive functional network; and 3) the characteristics of WM alterations related to EF at a whole brain level in PD-N, regardless of motor phenotype.

## 2. Patients and Methods

### 2.1. Subjects

PD patients consented and evaluated at the National Institute of Health (NIH, Bethesda, MD, USA) between 2011 and 2015 for a deep brain stimulation protocol conducted under Institutional Review Board-approved guidelines, were included in this retrospective study. All subjects had a diagnosis of PD according to the United Kingdom Parkinson’s Disease Society Brain Clinical Diagnosis Criteria [20,21]. The Mini Mental State Examination (MMSE) was used to define normal cognitive level [22]. The cutoff scores of MMSE were adjusted according to the educational level (> 20 for illiterates, > 25 for individuals with 1 to 4 years of education, > 26 for individuals with 5 to 8 years, > 28 for individuals with 9 to 11 years, > 29 for individuals with more than 11 years of education). The exclusion criteria were: 1) patients diagnosed with any neurological or psychiatric disease other than PD; 2) had global cognitive impairment (based on the Cognitive Impairment on Full Scale Intelligence Quotient (FSIQ) of WAIS-III Wechsler Adult Intelligence III, cut-off score less than 70); or 3) the use of illegal drugs. Cognitive function was evaluated in the clinically defined ON state by a certificated psychologist at the National Institute of Neurology and Stroke (NINDS, NIH, USA). All patients abstained from alcohol consumption for at least 24 hours before evaluations.

Physical and neurological evaluations were performed by NIH Parkinson’s Disease Clinic neurologists. The Unified Parkinson’s Disease Rating Scale part III (UPDRS-III) and the Hoehn and Yahr scale were used to each assess patient’s motor function [23,24]. The UPDRS-III was performed in the morning, after 12 hours off any dopaminergic medication (clinically defined OFF state). After the evaluation, patients took their regular dose of dopaminergic medications. One hour later, patients were tested for the UPDRS-III in the clinically defined ON state.

The total dosage of antiparkinsonian medication was converted to Levodopa equivalent dose (LED) according to an established method [25]. Motor phenotypes were derived from the UPDRS-III in the OFF state based on Jankovic’s classification system, as used in recent studies [4,26,27]. The tremor score was the sum of items 20 and 21, divided by 4. The PIGD score was the sum of items 22 to 27 and 31, divided by 15. Phenotypes were categorized by the ratio of each patients’ tremor score to his/her PIGD score. The TD-N group was defined as the ratio greater than or equal to 1.0. The PIGD-N group included all patients with a ratio less than or equal to 0.8. Subjects outside this range were excluded.

### 2.2. Neuropsychological tests

Neuropsychological tests were used to evaluate patients’ EF, including the Trail making test part A and B, the Stroop color naming, the Stroop word naming, the Stroop color-word interference task, as well as the FAS verbal fluency task and the animal category fluency task of Delis-Kaplan Executive Function System (D-KEFS) [28,29]. All the neuropsychological tests were assessed in the clinical ON state.

### 2.3. Image acquisition

MRIs from all patients were collected in the clinical ON state using a 3.0 T MRI scanner (Philips Achieva XT, Philips Medical Systems, Best, the Netherlands). The acquisition protocol included T1-weighted (T1w) turbo field echo, T2-weighted (T2w) turbo spin echo, and diffusion-weighted high-angle echo-planar imaging (DWI) sequences as we previous reported [30]. The T1w sequence was acquired with the following parameters: TR = 8.15 ms, TE = 3.735 ms, slice thickness = 1.00 mm, spacing between slices = 1.00 mm, echo train length = 240, FOV = 240 × 240 mm^2^, flip angle = 8°, acquisition matrix = 256 × 256 ×191, and total acquisition time = 6 min 53 s. The T2w sequence was collected with the following parameters: TR = 2500 ms, TE = 235.648 ms, slice thickness = 1.10 mm, spacing between slices = 0.55 mm, echo train length = 133, FOV = 250 × 250 mm^2^, flip angle = 90°, acquisition matrix = 256 × 256 x327, and total acquisition time = 4 min 37.5 s. The DWI sequence was collected with the following parameters: TR = 9776.51 ms, TE = 65 ms, slice thickness = 2 mm, spacing between slices = 2 mm, echo train length = 59, FOV = 224 × 224 mm^2^ (in-plane resolution 2 × 2 mm^2^), flip angle = 90°, acquisition matrix = 112 × 112 × 78, voxel size = 2 mm isotropic; 1 reference bo volume with no diffusion weighting (b=0 s/mm^2^) was collected, along with 33 non-collinear gradient directions with *b* = 1000 s/mm^2^. DWI acquisition time was 6 min 48 s.

### 2.4. Image processing

#### 2.4.1. Pre-processing

Image pre-processing was performed following our previously described method [30], utilizing the following software: Medical Image Processing, Analysis and Visualization (MIPAV) software package [31], TORTOISE (v. 3.1.2) [32], FreeSurfer (v. 5.0) [33], AFNI (v. 20.1.06) [34], SUMA [35] and FATCAT [36]. Briefly, anterior and posterior commissure (AC and PC) landmarks were manually defined on the mid-sagittal aligned T2w image using MIPAV. A rigid body transform was applied to T2w resulting in a horizontal AC-PC line and a vertical midsagittal plane. The re-aligned T2w images served as the co-registration target for the DWI volumes and the T1w volume.

The DWI volumes were first motion- and eddy-corrected (along with EPI distortion reduction), aligned to the target T2w volume and resampled to 1.5 mm isotropic voxels using TORTOISE’s ‘DIFFPREP’ program. The transformed volume was visually inspected. Uncertainty intervals of FA and principal diffusion directions were estimated with FATCAT’s ‘3dDWUncert’ using 500 jackknife resampling iterations, for subsequent use in probabilistic tractography, described below. The preprocessing script is included as Supplementary Material 1.

#### 2.4.2. ROIs definition and setup

FAS verbal fluency network: Gray matter (GM) regions of interest (ROIs) from the CA_ML_18_MNIA atlas in AFNI [37] were selected based on previous FAS verbal fluency fMRI studies [38,39] and include the bilateral prefrontal cortex pars triangularis and pars opercularis, inferior parietal lobule (IPL), inferior temporal gyrus (ITG), insula, middle temporal gyrus (MTG), superior parietal lobule (SPL), superior temporal gyrus (STG), thalamus, putamen, caudate, pallidum, and hippocampus (see Fig.1). Transformations were made using the AFNI’s ‘3dQwarp’ program [40]. Then, we nonlinearly mapped these ROIs to each subject diffusion space using AFNI’s ‘3dNwarpApply’ program, to be used as additional tractography targets within each subject’s native space. We estimated this network to include in our analysis because the FAS verbal fluency network was the only EF task showing significant differences between the two phenotypes in our study (see section 3.1).

**Fig. 1.**
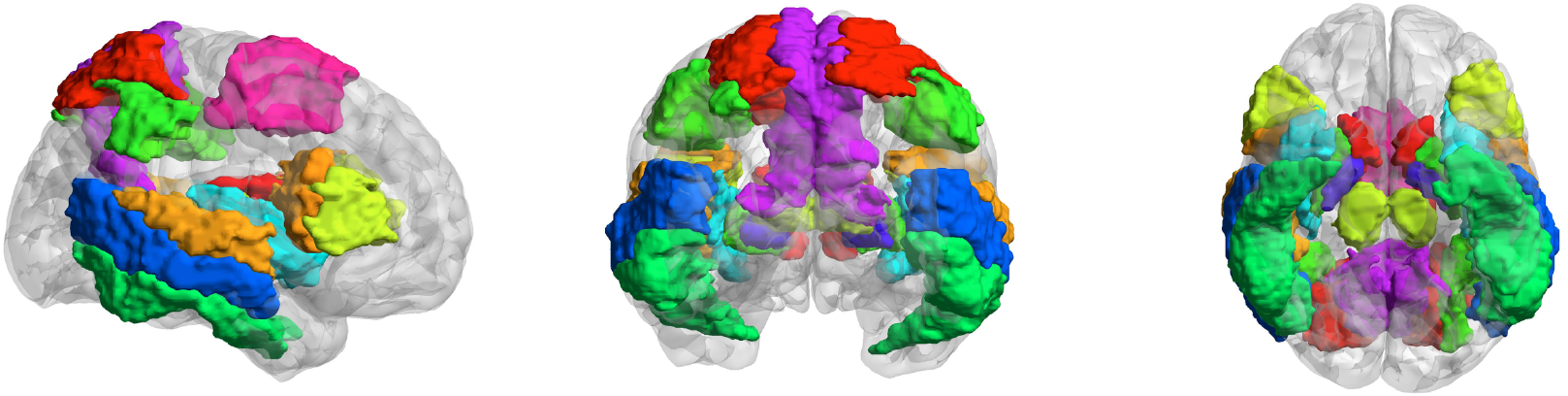
Target areas for tractography. 26 ROIs included in the FAS verbal fluency task-based network. Regions displayed in MNI space.

*Whole brain network:* In order to characterize the WM alterations related to EF at a whole brain level, regardless of motor phenotype, we used 83 GM ROIs from the Desikan-Killiany atlas of each subject’s FreeSurfer parcellation [37]. Additionally, the ROIs from the Talairach-Tournoux atlas (TT_Daemon) were identified to specify deep nuclei ROIs including bilateral red nucleus, substantia nigra, subthalamic nucleus, and hypothalamus. These deep nuclei were chosen based on prior reports related to cognitive functions, freezing gait, and other effects in PD indicating WM alterations [41-43]. The deep nuclei were added to GM ROIs by after transforming them to each individual’s diffusion data by AFNI’s ‘3dNwarpApply’. In total, 91 ROIs were included (detailed in Supplementary Material 2).

The procedures followed our previous reported method [30]. Briefly, the T1w volume was intensity-normalized using AFNI’s ‘3dUnifize’ and then skull stripped in FreeSurfer. The skull stripped T1w volume was registered to T2w volume using AFNI’s ‘align_epi_anat.py’ with the local Pearson’s coefficient (lpc) cost function [44]. The registered T1w was fed back into FreeSurfer for generating a cortical surface. Surface models and parcellation were created via another call to FreeSurfer’s ‘recon-all’ command. The definition of two networks and the wrapping processes of the two networks to subject’s space were described above. Each subject ROIs were inflated to the neighboring voxels surface and edges and stopped if it reached with another ROI (preventing ROI overlap) or voxels with an FA value larger than 0.2 (preventing WM overrun) using the FATCAT tool ‘3dROIMaker’, resulting in the two networks used for ROI-based probabilistic tractography.

#### 2.4.3. Probabilistic tractography

Probabilistic tractography was used to estimate the WM tracts from each pair of ROIs for the whole brain analysis and within the FAS verbal fluency network described above. In FATCAT, the probabilistic tractography analysis is based on the fiber assessment by continuous tracking including the diagonals (FACTID) algorithm, which has been shown to provide relatively robust results in both human and phantom testing studies [32]. We ran the program ‘3dTrackID’ in full probabilistic tracking mode (PROB), utilizing the tensor-uncertainty information described above, and the default tracking parameters (FA > 0.2, turning angle < 60, track length > 20 mm, 8 seeds per voxel, thresholding fraction > 0.021) with five seeds per voxel and a total of 5000 Monte Carlo iterations. Tracts passing through individual ROIs and locations of tracts that intersected any pair of ROIs were recorded. For each subject, an automatically generated ‘*.grid’ file output of ‘3dTrackID’ contained the statistical results describing the estimation of average DTI scalar parameters of interest (FA, MD, and RD) for WM ROIs connecting pairs of ROIs in the input GM network (91 × 91 for whole brain network, 26 × 26 for FAS verbal fluency network) in matrix format.

### 2.5. Statistics

#### 2.5.1. Clinical and neuropsychological tests analysis

AFNI’s 3dMVM and the R statistics software (v.3.6.2, R Foundation for Statistical Computing, Vienna, Austria) were used for statistical analysis [45]. Normality assumptions were evaluated based on the residuals using the Shapiro-Wilk test. Clinic and neuropsychological tests between PIGD-N and TD-N phenotypes were analyzed using repeated measures ANCOVA with age, disease duration, and education level as covariates. The categorical variables were calculated by fisher test. UPDRS-III ON and OFF scores were compared by paired-sample two-tailed *t*-test (*p* < 0.05).

#### 2.5.2. Effects of PD motor phenotypes on WM alterations related to EF

We analyzed WM differences between PIGD-N and TD-N phenotypes within the FAS verbal fluency network with an ANCOVA model, ran in AFNI’s ‘3dMVM’ program [45] (see Supplementary Material 3). Each subject’s age, disease duration years, and FAS verbal fluency task scores were set as covariates. FA, MD, and RD were defined as response variables, and the effect of each assessed in a separate model. Testing was carried out in a hierarchical manner. First, “network level” significance was tested with the omnibus F-statistic for the main effects of phenotype on the FAS verbal fluency network, as well as for the interactions between phenotype with FAS verbal fluency scores, age, and years of duration. Second, when the F-statistic was significant, post-hoc follow-up t-tests were used within the same 3dMVM command to investigate and pinpoint the “ROI level” significance of which WM connections had the strongest relationship driving the network-level effect. In other words, the network level F-statistic measured the primary effect of interest here, and when that showed a significant relation, then the ROI level post-hocs provided specific evidence about which parts of the network appeared to be driving the main effect (that is, which WM connections showed the strongest relationship, or whether the effect appeared to be relatively dispersed throughout the network). Due to the descriptive nature of the post-hoc tests, no multiple comparisons corrections were used. The omnibus F-test itself, a test of an intersection hypothesis, requires no multiple comparisons [46,47]

#### 2.5.3. Characteristics of WM alterations related to EF in PD-N

To estimate FAS verbal fluency scores effect on whole brain WM alterations, the ANCOVA model was separately used to estimate the FA, MD, and RD values in the whole brain network for the entire cohort (PD-N) regardless of phenotype (see Supplementary Material 3), following a similar strategy to the testing noted in the previous section for this whole brain network. In this part, we estimated the tractography at the whole brain level, as the FAS verbal fluency network is a part of the whole brain network. In the model, the FAS verbal fluency scores were defined as independent variables. DTI measurements were defined as separate response variables. FAS verbal fluency scores, age, and disease duration were included as covariates. The main effect of FAS scores was evaluated at the “network level” by omnibus F-statistics. When those showed significant relations, then post-hoc t-tests were performed to determine the most significant tracts “at the ROI level” between paired-ROIs contributing to the main effect.

## 3. Results

### 3.1. Demographics and neuropsychological tests

A total of 40 idiopathic PD subjects were recruited during the study period. Three subjects were excluded since they did not meet inclusion criterial (two had essential tremor and one had cervical dystonia). Another three subjects were excluded in the step of phenotype classifications. A total of 34 PD patients met all inclusion criteria and were included in the analysis. 19 patients were classified as TD-N and 15 as PIGD-N. The demographics and disease-related characteristics of included subjects are summarized in Table 1. EF scores of the entire PD cohort, and by the two phenotypes are described in Table 2. The FAS verbal fluency task showed significant difference between the two phenotype groups (*p* < 0.05, *t* _(32)_ = 2.230). The TD-N group showed a higher score (mean = 53.11, SD = 6.33) than the PIGD-N group (mean = 47.73, SD = 7.72). No other group differences were found. No significant correlations were found between neuropsychological results and UPDRS-III scores.

**Table 1.** Characteristics of the cohort of PD-N and two motor phenotypes.

|  | PD-N<br>(n=34) | TD-N<br>(n=19) | PIGD-N<br>(n=15) | *p* value<br>(TD-N vs.<br>PIGD-N) |
| --- | --- | --- | --- | --- |
| *Demographics* |  |  |  |  |
| Age (years) | 57.50 ± 7.79 | 57.26 ± 7.82 | 57.80 ± 8.03 | 0.846 |
| Gender (M/F) | 11M/ 23F | 5M/ 14F | 6M/ 9F | 0.701 |
| Education (years) | 15.31 ± 2.11 | 14.89 ± 1.94 | 15.80 ± 2.34 | 0.237 |
| *Disease-related symptoms* |  |  |  |  |
| MMSE | 29 ± 1.7 | 29 ± 2.4 | 29 ± 0.5 | 0.307 |
| Disease duration (years) | 12.79 ± 6.29 | 12.53 ± 5.63 | 13.13 ± 7.22 | 0.791 |
| LED (mg/day) | 901.30 ± 509.31 | 881.69 ± 554.36 | 926.80 ± 472.28 | 0.835 |
| UPDRS III OFF | 39.71±12.64 | 40.68 ±13.71 | 38.47 ±11.47 | 0.611 |
| UPDRS III ON | 33.23 ± 12.81 | 33.57 ±12.21 | 32.38 ± 13.42 | 0.831 |
| Tremor score | 1.85 ± 1.25 | 2.67 ± 1.03 | 0.82 ± 0.52 | < 0.001* |
| Rest tremor | 0.45 ± 0.19 | 0.39 ± 0.19 | 0.53 ± 0.16 | 0.026* |
| Active tremor | 0.22 ± 0.10 | 0.19 ± 0.10 | 0.27 ± 0.08 | 0.014* |
| PIGD score | 1.70 ± 0.60 | 1.61 ± 0.60 | 1.89 ±0.54 | 0.163 |
| Rigidity score | 1.35 ± 0.89 | 1.14 ± 0.85 | 1.63 ± 0.91 | 0.118 |
| Bradykinesia score | 2.23 ± 0.95 | 1.95 ± 0.97 | 2.67 ± 0.82 | 0.025* |
| DRS-II | 138.74 ± 4.85 | 138.58 ± 5.14 | 138.93 ± 3.94 | 0.823 |
| BDI | 7.27 ± 5.37 | 8.63 ± 5.97 | 5.53 ± 4.03 | 0.081 |
| FSIQ | 101.7 ± 13.15 | 104.63 ± 14.42 | 98.6 ± 11.3 | 0.181 |

Note: \* statistically significant ( $p < 0.05$ ). MMSE = the Mini-Mental State Examination. LED = Levodopa dosage equivalence. UPDRS-III = Unified Parkinson's Disease Rating Scale part III. DRS-II = Dementia rating scale-II. BDI = Beck Depression Inventory. FSIQ = Full Scale Intelligence Quotient. The following motor scores were from items in UPDRS-III. Tremor score = the sum of items 20 and 21, divided by 4. Rest tremor score = the sum of item 20 divided by 5. Active tremor score = the sum of item 21 divided by 2. PIGD
score = the sum of items 22-27 and 31, divided by 15 from. Rigidity score = the sum of item 22 divided by 5. Bradykinesia score = item 23.

**Table 2.** Executive Function comparison across motor phenotypes.

|  | PD-N<br>(n=34) | TD-N<br>(n=19) | PIGD-N<br>(n=15) | *p* value<br>(TD-N<br>vs.<br>PIGD-N) |
| --- | --- | --- | --- | --- |
| Trail Making A | 48.06 ± 11.02 | 50.05 ± 10.92 | 45.43 ± 10.99 | 0.231 |
| Trail Making B | 47.85 ± 8.67 | 46.44 ± 11.92 | 45.93 ± 9.15 | 0.889 |
| Stroop Word Reading | 44.94 ± 7.52 | 43.11 ± 8.23 | 47.27 ± 5.98 | 0.098 |
| Stroop Color Naming | 41.68 ± 8.07 | 42.05 ± 7.20 | 41.2 ± 9.29 | 0.773 |
| Stroop Color-Word Interference | 48.91 ± 9.02 | 48.68 ± 10.48 | 49.2 ± 7.63 | 0.869 |
| FAS Verbal Fluency | 51.41 ± 11.22 | 53.11 ± 6.33 | 47.73 ± 7.72 | 0.033* |
| Animal Verbal Fluency | 49.50 ± 9.56 | 48.21 ± 7.9 | 49.47 ± 10.34 | 0.699 |

Note: \* statistically significant ( $p < 0.05$ ). Values was expressed as mean ± standard deviation. All subtests of EF are from Delis-Kaplan Executive Function System.

### 3.2. Effects of PD motor phenotypes on alterations of WM related to EF

ANCOVA results showed significant difference between the two phenotypes in the MD and RD of the WM connecting task-based ROIs after controlling for age, disease duration, and scores of the FAS verbal fluency task [F_RD (1,18)_ = 12.62, *p* = 0.0004. F_MD (1,18)_ =14.26, *p* = 0.0002]. Post-hoc *t*-tests indicated significant differences in MD and RD in the left SLF, connecting the left STG and left insula, and in the right fronto-insular fibers, anatomically connecting the pars opercularis and insula [48]. The right frontal U-fibers connecting pars opercularis and pars triangularis, and the right temporal U-fibers connecting ITG and MTG showed statistically significant differences in the post-hoc analysis of RD (described in Table 3). Phenotype × FAS scores interactions were significant for MD and RD [F_RD (1,18)_ = 4.262, *p* = 0.039; F_MD (1, 18)_ = 5.20, *p* = 0.023]. Neither an interaction in phenotype × age nor in phenotype × disease duration was found.

**Table 3.** FAS verbal fluency network: MD and RD results.

| ROIs pair | anatomical WM fibers | TD-N | PIGD-N | *p* value |
| --- | --- | --- | --- | --- |
| MD ($10^{-3}$ mm<sup>2</sup>/s) | | | | |
| lh-superior temporal gyrus__lh-insula | left superior longitudinal fasciculi | $0.775 \pm 0.023$ | $0.758 \pm 0.040$ | 0.005 |
| rh-pars opercularis gyrus__rh-insula | right fronto-insular tracts | $0.740 \pm 0.022$ | $0.750 \pm 0.031$ | 0.004 |
| RD ($10^{-3}$ mm<sup>2</sup>/s) | | | | |
| lh-superior temporal gyrus__lh-insula | left superior longitudinal fasciculi | $0.603 \pm 0.024$ | $0.605 \pm 0.036$ | 0.005 |
| rh-inferior temporal gyrus__rh-middle temporal gyrus | right temporal U-fibers | $0.602 \pm 0.019$ | $0.599 \pm 0.029$ | 0.01 |
| rh-pars opercularis__rh-insula | right fronto-insular tracts | $0.589 \pm 0.025$ | $0.598 \pm 0.035$ | 0.003 |
| rh-pars opercularis__rh-pars triangularis | right frontal U-fibers | $0.588 \pm 0.024$ | $0.607 \pm 0.064$ | 0.01 |

Note: values are expressed as mean ± standard deviation. rh = right hemisphere. lh = left hemisphere. The ANCOVA was performed with subjects' age, years of disease duration, FAS verbal fluency task scores as covariates.

### 3.3. Characteristics of WM integrity correlated with performance of FAS verbal fluency task in the cohort of PD-N patients

The ANCOVA model showed significantly increased MD in tracts connecting cortico-cortical and cortico-subcortical regions in both hemispheres [F _(1, 31)_ = 4.89, *p* = 0.027], but not significant FA or RD differences were found (detailed in Table 4).

**Table 4.** Effects of FAS verbal fluency scores in the whole brain network for the PD-N cohort: MD results.

| ROIs pair | MD ($10^{-3}$ mm<sup>2</sup>/s) | *p* value |
| --- | --- | --- |
| Cortical regions |  |  |
| Frontal cortex |  |  |
| rh-caudal middle frontal gyrus__rh-superior frontal gyrus | $0.737 \pm 0.030$ | 0.011 |
| rh-caudal middle frontal gyrus__rh-precentral cortex | $0.730 \pm 0.025$ | 0.015 |
| rh-pars triangularis__ rh-rostral middle frontal gyrus | 0.757 ± 0.053 | 0.034 |
| rh-medial orbitofrontal cortex__ rh-rostral anterior cingulate cortex | 0.804 ± 0.038 | 0.024 |
| Temporal cortex |  |  |
| rh-inferior temporal gyrus__ rh-insula | 0.781 ± 0.031 | 0.045 |
| rh-inferior temporal gyrus__ rh-superior temporal gyrus | 0.768 ± 0.026 | 0.036 |
| Parietal cortex |  |  |
| rh-superior parietal cortex__ rh-supramarginal gyrus | 0.767 ± 0.037 | 0.037 |
| Occipital cortex |  |  |
| lh-cuneus cortex__ lh-pericalcarine cortex | 0.780 ± 0.040 | 0.028 |
| Fronto-temporal cortex |  |  |
| rh-pars triangularis__ rh-insula | 0.745 ± 0.024 | 0.018 |
| rh-precentral cortex__ rh-insula | 0.742 ± 0.030 | 0.012 |
| Fronto-parietal cortex |  |  |
| rh-postcentral cortex__ rh-precentral cortex | 0.753 ± 0.030 | 0.023 |
| lh-precentral cortex__ lh-superior parietal cortex | 0.734 ± 0.028 | 0.037 |
| Temporo-parietal cortex |  |  |
| rh-postcentral cortex__ rh-insula | 0.76 ± 0.0370 | 0.048 |
| rh-supramarginal gyrus__ rh-insula | 0.756 ± 0.031 | 0.042 |
| Parieto-occipital cortex |  |  |
| lh-pericalcarine cortex__ lh-precuneus cortex | 0.805 ± 0.041 | 0.047 |
| rh-cuneus cortex__ rh-precuneus cortex | 0.765 ± 0.035 | 0.021 |
| rh-fusiform gyrus__ rh-para hippocampal gyrus | 0.832 ± 0.115 | 0.049 |
| Cortico-subcortical regions |  |  |
| lh-Pallidum__ lh-insula cortex | 0.731 ± 0.051 | 0.044 |

Note: values are expressed as mean ± standard deviation. rh: right hemisphere. lh: left hemisphere. Age, years of duration, and phenotype were defined as covariates in the ANCOVA model.

## 4. Discussion

In this study, we investigated the differences in WM integrity underlying the FAS verbal fluency network between the PIGD-N and the TD-N phenotypes, and analyzed the WM properties related to EF in PD-N patients at the whole brain level, regardless of phenotype effect. Our results showed a greater loss in WM integrity with lower EF in PIGD-N compared to TD-N phenotype, which is in agreement with a previous study [14]. In addition, we confirmed that SLF MD and RD values are correlated with EF in PD-N patients, as previously reported [12,49–52].

### 4.1. Cognitive results

Our PIGD-N cohort had lower scores in the FAS verbal fluency task than the TD-N one, which is in agreement with a previous study [12]. Other studies reported that patients with the PIGD-N phenotype had lower scores in other cognitive tasks, such as MMSE, Montreal Cognitive Assessment, Attention, and the Trail Making Tests when compared to patients with the TD-N phenotype [6,53,54], suggesting that global cognitive functions differs between the two phenotypes, even when cognition is considered normal. The neuropsychological tests included in our study are closely related with multiple neuropsychological processes of EF. The processes underlying the Trail Making A and B tests demanded high graphomotor ability [55]. The Stroop color-word interference and verbal fluency tasks demanded high working memory, association, selection and inhibition abilities [56–58]. We found no significant differences between the two phenotypes for the Trial Making Tests in our study. These suggests that tasks such as association and selection were more fragile than graphomotor abilities in the PIGD-N group, or both subgroups were equally affected.

### 4.2. FAS verbal fluency network and PD motor phenotype

The WM tract connecting STG with insula showed increased MD and RD in the PIGD-N group. The tract was anatomically matched to the SLF, an important tract for EF. Previous studies reported decreased FA in SLF and corpus callosum in PIGD compared to normal control, but no such differences were found in TD [14,49,59–62]. These previous results used different processing methods, did not account for cognitive function, and compared PIGD-N to normal controls, rather than between PD motor phenotypes. Therefore, due to methodological differences, our results are not directly comparable with those studies. However, as a collection, these studies all point towards the presence of an altered SLF in the PIGD-N group.

In addition, the WM tracts between ROI pairs, including the IFG pars opercularis gyrus and insula, pars opercularis and pars triangularis, right ITG with MTG, showed significant RD differences between the two phenotypes. The pars opercularis and insula are reportedly involved in speech articulation, vocalization of emotional states, and facial expression [48]. Especially, the WM tracts between pars opercularis and pars triangularis are involved in semantic processing, including lexis association and selection [63]. Thus, our results suggest that both facial muscle rigidity and language processing might be altered in PIGD-N phenotype. Compared to the TD phenotype, increased MD in tracts among the right inferior parietal lobe, premotor, and primary motor cortex in PIGD phenotype were previously reported [59]. These regions were not included in our FAS verbal fluency network, and at the whole brain level we did not compare phenotypes, as we focused on the effect of FAS verbal fluency in PD-N irrespective of phenotype. Therefore, we cannot compare those findings to our results.

### 4.3. Characteristics of WM integrity correlated with performance of FAS verbal fluency task in the cohort of PD-N patients

In our cohort of PD-N patients, the WM alterations related to the FAS verbal fluency task scores were characterized by increases in MD of inter- and intra-regional connections between cortex and subcortex in both hemispheres. In the frontal cortex we observed MD changes in the short U tracts between the right caudal MFG and the SFG, and in the U tracts connecting the right MFG and the precentral cortex. The function of these connections are still not clear, but the three regions are connected with the striatum through descending projection fibers [48]. The connections between medial orbitofrontal cortex and anterior cingulate cortex are part of the fronto-orbitopolar tract [64,65]. The tracts connecting the left precentral cortex and superior parietal cortex are important in attention and spatial visual abilities [66]. Duncan et al. reported significant decreased phonemic and semantic fluency scores in PD-N compared to normal controls [19]. They reported increased MD in connections between frontal and parietal WM which correlated with deficits in semantic fluency performance in PD-N. Although we evaluated a different cognitive task, our observed alterations in tracts connecting frontal and parietal cortex are in line with their findings. It is possible that both phonemic and semantic verbal fluency tasks share basic circuits for lexis association and selection processes [67]. We reported alterations of tracts connecting right frontal and insular regions as well as the tracts connecting right frontal and striatal regions. Fronto-insular-striatal tracts are involved in attention, sensory memory, and motor speech functions, suggesting these cognitive domains are disturbed in PD-N patients [68,69]. We also found the SLF integrity was correlated to the EF in PD, as previous studies reported [12,49–52].

We found alterations in MD, but not in FA and RD, in the cohort of PD-N patients, in agreement with previous studies [12,19]. This suggests that PD-N might present changes related to membrane integrity, which could be caused by cellularity, edema, or necrosis [70]. In addition, we reported the characteristic parameters related to EF in PIGD-N are the increased MD and RD, not the FA. RD changes have been associated with loss of the integrity and properties of the myelin sheath [71]. Other studies reported a significant reduction in FA in the left parietal, and prefrontal WM in non-demented PD patients with EF impairment tested by Wisconsin card sorting test [72-74]. These studies were not restricted to PD-N, and different neuropsychological tests may result in the differences. Thus, our results suggest MD and RD values could aid detecting WM alterations related to EF in PD patients having normal cognitive functions, especially for the PIGD phenotype.

There are some limitations of this study. We acknowledge a small sample set per subgroup, limiting our results. We compared cognitive scores using a cross-sectional design, longitudinal studies could shed more light into the WM integrity changes and their relationship with behavior. Tractography is not a perfect tool for mapping WM pathways, resulting in both false “negatives” and “positives” in its connection maps; however, it provides a methodology for parcellating the WM skeleton relatively consistently across a group, using each subject’s own data. Considering the motor phenotype can shift from TD to PIGD phenotype, the repeated prospective measurements in a longitudinal design might be of interest. Finally, we did not examined the medication effect on behavior.

## 5. Conclusion

The present study reinforces the concept of overt clinical cognitive deficits present in PD-N. PIGD-N shows more microstructural changes related to when compared to TD-N phenotype. The mechanisms of phenotype effects on WM integrity alterations related to cognitive functions can be comprehensively studied in longitudinal studies in the future.

## Data Availability

The data referred to this study is not avaiable to use regarding the confidentiality agreement.

## Acknowledgments and Funding

This work was supported by the NINDS Intramural Research Program; and the National Institutes of Health Graduate Partnerships Program. QY stipends in NIH was supported by the Joint-Ph.D. program of the China Scholarship Council. PAT was supported by the NIMH and NINDS Intramural Research Programs (ZICMH002888) of the NIH (HHS, USA). We sincerely thank Dr. Gang Chen for his advice on statistics.

